# Evaluation of commercially available immuno-magnetic agglutination and enzyme-linked immunosorbent assays for rapid point-of-care diagnostics of COVID-19

**DOI:** 10.1101/2020.08.15.20172080

**Authors:** Maria Engel Moeller, Jeppe Fock, Pearlyn Pah, Antia De La Campa Veras, Melanie Bade, Marco Donolato, Simone Bastrup Israelsen, Jesper Eugen-Olsen, Thomas Benfield, Frederik Neess Engsig

## Abstract

**Introduction:** Coronavirus Disease 2019 (COVID-19) is caused by *severe acute respiratory coronavirus-2* (SARS-CoV-2). Fast, accurate and simple blood-based assays for quantification of anti-SARS-CoV-2 antibodies are urgently needed to identify infected individuals and keep track of the spread of disease.

**Methods:** The study included 35 plasma samples from 22 individuals with confirmed COVID-19 by real time reverse-transcriptase-polymerase-chain-reaction and 40 non-COVID-19 plasma samples. Anti-SARS-CoV-2 IgM/IgA or IgG antibodies were detected by a microfluidic quantitative immunomagnetic assay (IMA) (ViroTrack Sero COVID IgM+IgA /IgG Ab, Blusense Diagnostics, Denmark) and by enzyme-linked immunosorbent assay ((ELISA) (EuroImmun Medizinische Labordiagnostika, Germany).

**Results:** Of the 35 plasma samples from the COVID-19 patients, 29 (82.9%) were positive for IgA/IgM or IgG by IMA and 29 samples (82.9%) were positive by ELISA. Sensitivity for only one sample per patient was 68% for IgA+IgM and 73% IgG by IMA and 73% by ELISA. For samples collected 14 days after symptom onset, the sensitivity of both IMA and ELISA was around 90%. Specificity of the IMA reached 100% compared to 95% for ELISA IgA and 97.5% for ELISA IgG.

**Conclusion:** IMA for COVID-19 is a rapid simple-to-use point of care test with sensitivity and specificity similar to a commercial ELISA.

## INTRODUCTION

Coronavirus disease 2019 (COVID-19) is caused by severe acute respiratory syndrome coronavirus 2 (SARS-CoV-2) and has spread globally since its discovery in Wuhan, China in December 2019 [1,2]. In spite of advances in antiviral treatment it remains a disease with considerable morbidity and mortality [3,4],

Real time reverse-transcription polymerase chain reaction (RT-PCR) detection of SARS-CoV-2 RNA is the recommended test to diagnose active COVID-19 but several serological tests for COVID-19 have been developed [5-8]. Immunoassays detect different antibodies to SARS-CoV-2, namely antibodies to different parts of the spike or the nucleocapside protein [9-12], While serological testing in general cannot replace RT-PCR for diagnosing acute COVID-19, it may serve as a valuable supplement in persons with classical symptoms of COVID-19 and repeated negative RT-PCR, although its main application is to assess immunity.

Enzyme-linked immunosorbent assay (ELISA) tests may take hours to perform, are usually batched and require laboratory facilities and skilled personnel. Lateral flow assays for antibody detection are quick single sample tests, but have lower sensitivity compared to ELISA, the read-out is operator dependent, and the result is qualitative [13-15]. An automated, real-time and quantitative point-of-care test using capillary blood with high sensitivity would offer the ability of testing for SARS-CoV-2 antibodies both within and outside of a hospital setting.

In this study, we compared the performance of a well-tested commercial ELISA for COVID-19 with a newly developed automated immunomagnetic assay (IMA) technology for rapid testing for COVID-19 antibodies.

## MATERIALS AND METHODS

### Subjects and samples

We included individuals with confirmed COVID-19 by RT-PCR for SARS-CoV-2 RNA on naso-/oropharyngeal swabs or lower respiratory tract specimens, from March 20 to May 1, 2020 with at least one available plasma samples[16]. Plasma samples collected prior to July 2019 from a biobank for Danish HIV infected individuals served as COVID-19 negative controls [17]. Samples were stored at −80 °Celsius until testing. Demographic and clinical data on the individuals were transferred from electronic health records. A waiver of individual informed consent was granted by the Regional Ethics Committee of the Capital Region of Denmark (record no. H-20040649). The study was further approved by the Danish Patient Safety Authority (record no. 31-1521-309) and the Regional Data Protection Center (record no. P-2020-260). Data was entered into an electronic data capture tool hosted by the Capital Region of Denmark [18,19]. Variables included age, gender, comorbidity, radiographic findings, duration of symptoms, supplemental oxygen, do not resuscitate orders, intensive care, mechanical ventilation and 30-day mortality. In this paper, severe disease was defined as need of more than 15 liters of supplementary oxygen per minute.

Blinded samples were measured by ViroTrack Sero Covid IgA+M/IgG (Blusense Diagnostics, Denmark) (IMA) and EI ELISA IgA/IgG (EuroImmun Medizinische Labordiagnostika, Germany) (ELISA) as described below.

### ViroTrack Sero Covid IgM+lgA /IgG

In brief, 10 ul of plasma was mixed with 150 ul sample dilution buffer, vortexed and 50 ul of the diluted plasma was loaded on to the microfluidic cartridge. The IMA IgM+IgA/IgG antibody (Ab) test utilizes a centrifugal microfluid platform together with optomagnetic readout based on the agglutination of magnetic nanoparticles (IMA). The sample was manipulated on a cartridge with the help of the centrifugal force, Coriolis force and Euler force to allow for separation, sedimentation, aliquoting, and reagent re-suspension by the design of microfluidic chambers and channels and control over the angular velocity profile of the cartridge rotation [20]. The optomagnetic signal was obtained by measuring the modulated transmitted light through a suspension of magnetic nanoparticles in response to an alternating magnetic field [21]. The magnetic particles were covalently coupled to antigens or antibodies. Upon target induced magnetic particle agglutination the change in optical and magnetic anisotropy results in a change in the optomagnetic signal which can be used to quantify the target concentration [22-24], Incubating the particle in homogeneous magnetic fields speeds up the reaction kinetics [22,25]. IMA does not require a secondary antibodies. The magnetic particles were functionalized with SARS-CoV-2 nucleocapsid recombinant protein in the IMA IgM+IgA/IgG Ab kit. Negative results were reported with values below 3.5 IMA units and positive results with 4.5 IMA and above. The equivocal region (borderline results) is between 3.5 and 4.5 IMA. The cutoff value of 4.5 IMA was determined from multiple measurements on negative samples to ensure less than 0.1% false positives with 95 % confidence. Values above 20 units were classified as high.

### Euroimmun ELISA (IgA, IgG)

ELISA IgA/IgG was performed according to the manufacturer’s specifications. In both ELISAs the antigen used is glycosylated Spike 1 protein. 1.2 μl of plasma was diluted in 118.8 I sample buffer provided in the kit. 110 μl of the sample was first transferred to an uncoated 96 well plate, and subsequently transferred to a coated 96 well ELISA plate using an eighth channel pipette and incubated at 37 °C for 1 h. The wells were emtied and subsequently washed three times with wash buffer provided in the kit. 100 μl of conjugate solution was added to each well, and the plate incubated at 37 °C for 30 min, and subsequently washed three times with wash buffer. 100 μl of substrate was added to each well and the plate incubated in dark at room temperature for 30 min. 100 μl of stop-solution was added and the ELISA plate was measured in a Multiskan FC Microplate Photometer (Thermo Scientific, type 357) at 450 nm. The results were evaluated in terms of the absorbance ratio of the sample and a calibrator. Negative results were reported with ratio below 0.8 and positive results with ratios above 1.1 and above. The equivocal region (borderline results) is between 0.8 and 1.1.

### Statistics

Patient characteristics were presented as median with interquartile range or count with percentage. The data analysis included calculation of the following parameters: sensitivity, specificity, positive predictive value (PPV), negative predictive value (NPV), and accuracy. Confidence intervals for sensitivity and specificity were calculated using the Exact Binomial method [26]. As case samples were taken from individuals identified as having COVID-19 from a positive RT-PCR, these were considered ‘true positive’. The plots including receiver operating characteristics (ROC)-curves were constructed in python using the matplotlib, seaborn and sklearn packages. Differences in titers were calculated by Fisher’s exact test using SPSS statistical software, Version 25.0 (Norusis; SPSS Inc., Chicago, Illinois, USA).

## RESULTS

We included a total of 22 individuals, who contributed 35 plasma samples. The individuals were mostly male (63,6%) with a median age of 72 years of whom most had at least one co-morbidity (83,6%). All had chest radiograph infiltration, and of these, 73% had multilobular infiltration. Forty plasma samples from Danish HIV infected individuals collected prior to July 2019 were included as controls.

Twenty-one received supplementary oxygen; maximum support during admission was 0-14 liters: four persons (18%), 15-29 liters; five persons (23%) and > 30 liters; 13 persons (59%). 41% of the patients were labeled “do-not-resuscitate” (DNR) and 29% were limited in treatment in terms of intensive care unit (ICU) admission. 41% of the patients were admitted to the ICU. 18% of the patients died within 30 days. (Table S1)

Five individuals were sampled over multiple days (POS 1-5). Figure 1 shows the evolution of the obtained results from the ELISA and IMA. For patient POS 1 and POS 2 we observed a change from negative at day 23 and 10, respectively, to stable positive for all subsequent samples by both assays (ELISA IgA and IMA IgA+IgM). Similarly, levels of IgG turned positive. POS 3, 4 and 5 were stable too, increasing over time. POS 1 (day 37) showed a decrease of IgG levels over time by ELISA and IMA. Of the 18 patients with severe disease, 13 (72%) had high IgM+A and/or IgG IMA titers versus 1 (25%) among the four patients with non-severe disease (p= 0.117) (Table S1).

**Figure 1:**
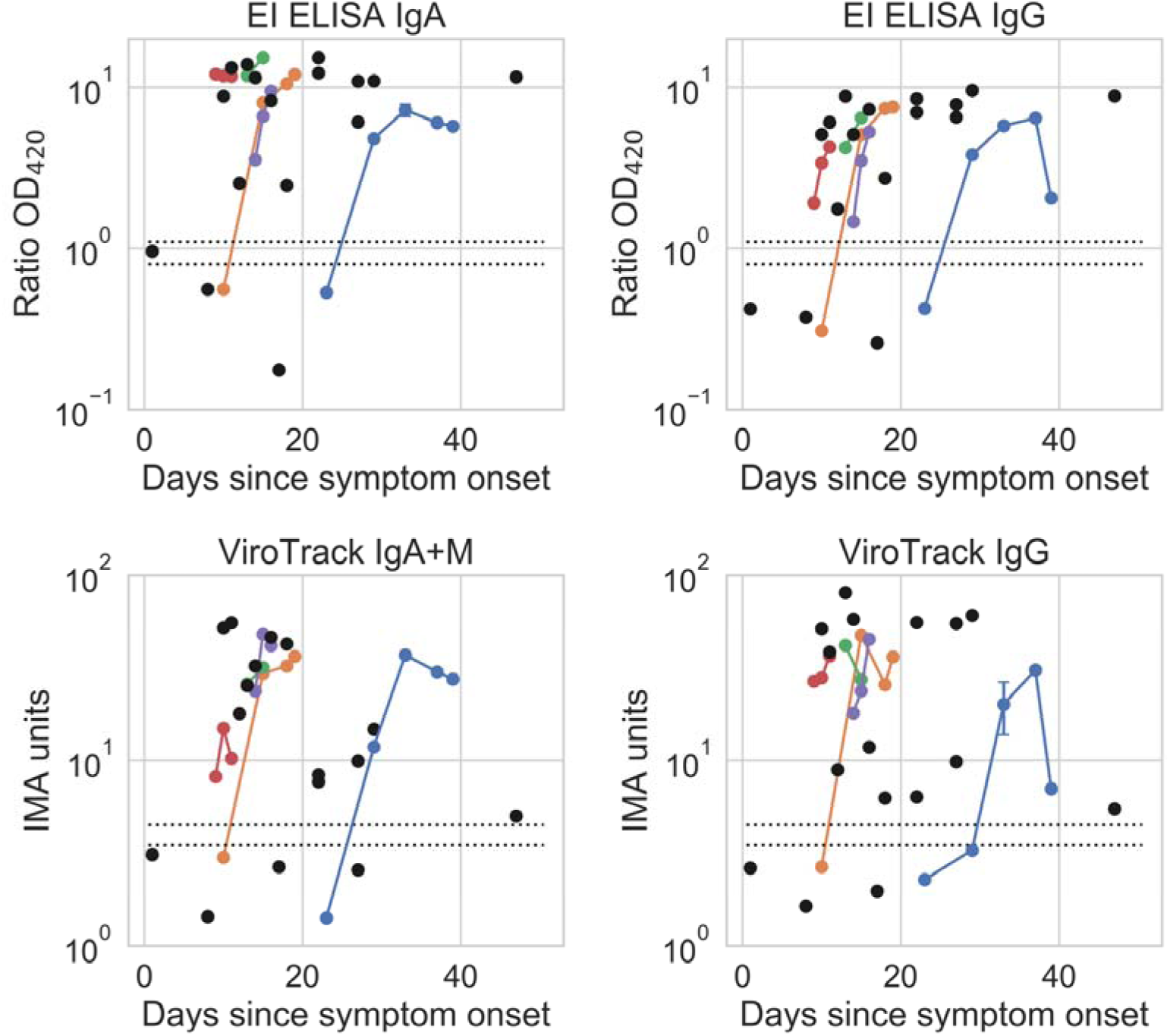
Severe acute respiratory coronavirus-2 antibody titer versus days after symptom onset. One samples, measured negative or borderline by microfluidic quantitative immunomagnetic assay (IMA) (Virotrack) and enzyme-linked immunosorbent assay (ELISA), did not have information on the days since symptom onset and are not included in the plot. Black points are from patients with only one sample, and colored are from POS 01 (blue), POS 02 (yellow), POS 03 (green), POS 04 (red) and POS 05 (purple). The dotted lines indicate the cut-off values to determine positive and negative test results and the zone in between represents the borderline results. El: Euroimmun.

### Assay performance

#### Sensitivity Analysis

Out of the 35 positive RT-PCR SARS-CoV-2 samples (all samples, single and longitudinal, included), 28 were IgA+IgM positive and 28 were IgG positive by IMA, and 29 positive for IgA and IgG by ELISAs (Table 1). This corresponds to a sensitivity of 80.0% for IgA+IgM and IgG by IMA and to 82.9% by ELISA. By combining the IMA IgA+IgM and IgG and the ELISA IgA and IgG results, the sensitivity was comparable for both assays (82,9%).

**Table 1:** Analytical sensitivity, specificity, and predictive values for the detection tests, considering all 35 samples from the 22 individual patients as positive samples. Equivocal/borderline results for enzyme-linked immunosorbent assay (ELISA) were treated as negatives (eqv. Neg) and as positives (eqv. Pos). No equivocal/borderline results were found in the microfluidic quantitative immunomagnetic assays (IMA) (Virotrack). The confidence intervals of the analytical sensitivities, specificities, and predictive values are given in table S2. El: Euroimmun.

|  |  | Sensitivity | Specificity | Agreement | PPV | NPV |
| --- | --- | --- | --- | --- | --- | --- |
| EI ELISA<br>(eqv. Neg) | IgA | 29/35 (82.9%) | 38/40 (95.0%) | 67/75 (89.3%) | 29/31 (93.5%) | 38/44 (86.4%) |
|  | IgG | 29/35 (82.9%) | 39/40 (97.5%) | 68/75 (90.7%) | 29/30 (96.7%) | 39/45 (86.7%) |
|  | IgA+IgG | 29/35 (82.9%) | 38/40 (95.0%) | 67/75 (89.3%) | 29/31 (93.5%) | 38/44 (86.4%) |
| EI ELISA<br>(eqv. Pos) | IgA | 30/35 (85.7%) | 35/40 (87.5%) | 65/75 (86.7%) | 30/35 (85.7%) | 35/40 (87.5%) |
|  | IgG | 29/35 (82.9%) | 39/40 (97.5%) | 68/75 (90.7%) | 29/30 (96.7%) | 39/45 (86.7%) |
|  | IgA+IgG | 30/35 (85.7%) | 35/40 (87.5%) | 65/75 (86.7%) | 30/35 (85.7%) | 35/40 (87.5%) |
| Virotrack | IgA+IgM | 28/35 (80.0%) | 40/40 (100%) | 68/75 (90.7%) | 28/28 (100%) | 40/47 (85.1%) |
|  | IgG | 28/35 (80.0%) | 40/40 (100%) | 68/75 (90.7%) | 28/28 (100%) | 40/47 (85.1%) |
|  | IgA+IgM+IgG | 29/35 (82.9%) | 40/40 (100%) | 69/75 (92.0%) | 29/29 (100%) | 40/46 (87.0%) |

None of the IMA readings were borderline, whereas one of the 35 positive samples was borderline positive for IgA by ELISA (absorbance ratio between 0.8 and 1.1). Including the equivocal result as a positive increased the sensitivity for ELISA IgA to 85.7% (Table 1). The positive predictive value (PPV) of all the methods used here (ELISA and IMA) is likely to be 93.5% or higher, due to the high specificities observed (ranging from 95.0 to 100%).

By considering the first drawn samples from each individual, the sensitivity was 68% for IgA+IgM and 73% for IgG by IMA and 73% by ELISA. By combining the IMA IgA+IgM and IgG and the ELISA IgA and IgG results, the sensitivity was comparable for both assays (73%). Days from the first symtom to drawing of the first sample ranged from 1-47 days, with a median of 14 days. For samples collected 14 days after sympthoms onset the sensitivity of both IMA and ELISA test was around 90%.

### Specificity Analysis

The specificity of IMA was 100.0% for the IgM+IgA and the IgG assays, respectively. Three of the 40 controls samples (two IgA and one IgG) were positive by ELISA resulting in a specificity of 95.0% and 97.5%, respectively (Table 1). Additionally, three samples were borderline positive by ELISA for IgA corresponding to a lower specificity of 87.5% (Table S1). No sample was indeterminant by IMA.

### Semi-quantitative results of IgA, IgM and IgG detection

The distance of data points from the cut-off values and confidence in assigning a positive or negative result differed between the IMA and ELISA assays (Figure 2). The distribution of positive and negative data points was distinct for IMA cartridge, with a cut-off value above all the negative samples, which allowed for unequivocal interpretation of all measurements. In contrast, the ELISA data had less separation, especially for IgA, resulting in a ‘grey zone’ of borderline data points to which a positive or negative result could not be assigned. Both positive and negative samples have borderline data points. Receiver operating characteristic (ROC) curves of the assays all showed area under curve above 0.91 (see Figure S1).

**Figure 2:**
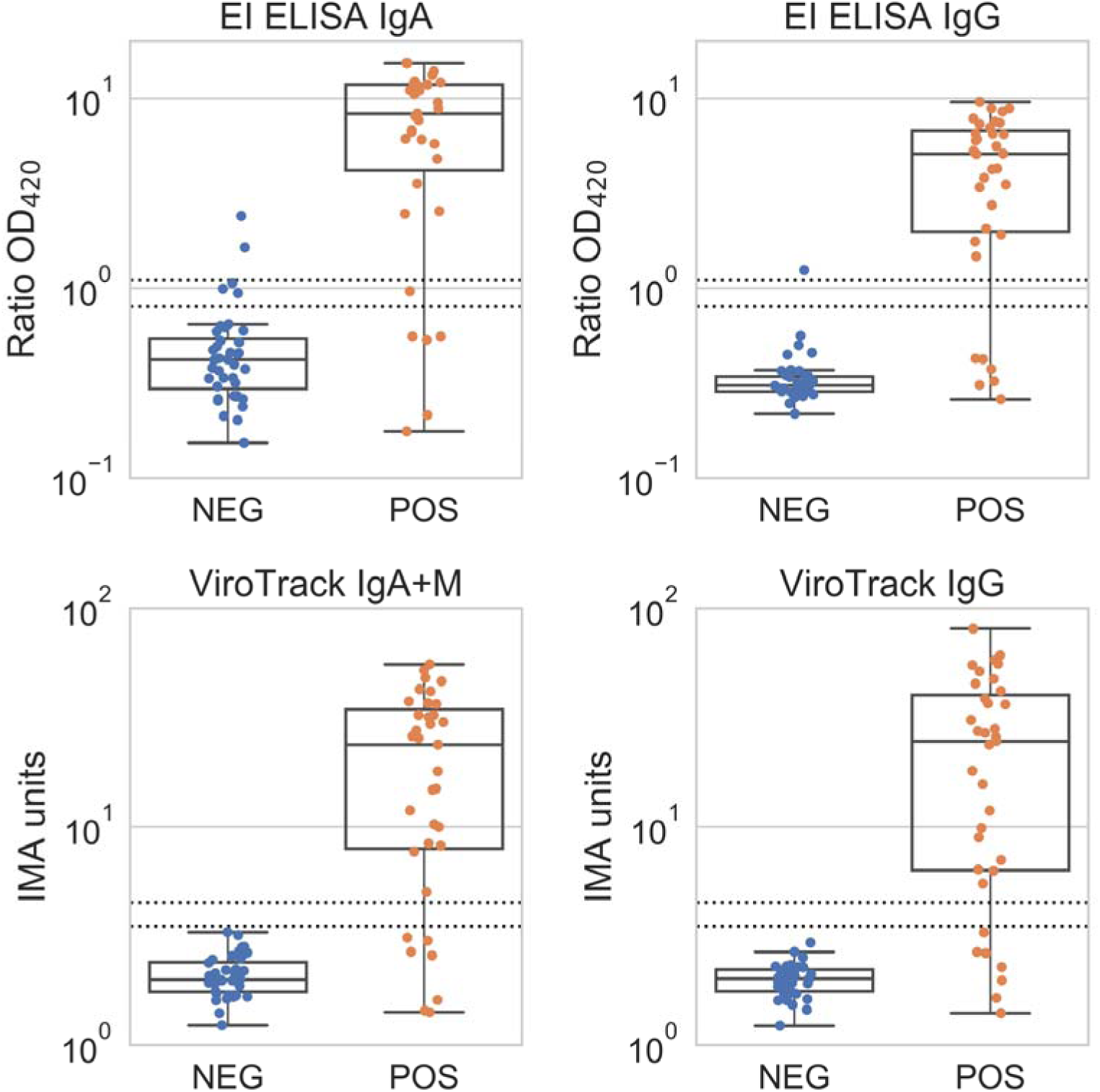
Severe acute respiratory coronavirus-2 antibody assay performance. Data distribution obtained for positive (POS) and negative (NEG) samples, confirmed by real time reverse-transcriptase-polymerase-chain-reaction using (A) enzyme-linked immunosorbent assay (ELISA) IgA and IgG, and (B) microfluidic quantitative immunomagnetic assay (IMA) (Virotrack) IgM+IgA and IgG. Lines represent median values with minimum to maximum ranges. The dotted lines indicate the cut-off values to determine positive and negative test results, the zone in between represents the borderline results. El: Euroimmun.

## DISCUSSION

This study showed that IMA IgM+IgA/IgG performed similar to a commercial ELISA with a sensitivity for each assay of 80% and a combined sensitivity of 82,9% to detect antibodies to SARS-CoV-2 in patients with moderate to severe disease. Sensitivity for only one sample per patient (first sample drawn) was 68% for IgA+IgM and 73% IgG by IMA and 73% by ELISA. For samples collected 14 days after symptom onset, the sensitivity of both IMA and ELISA was around 90%. The specificity was 100% by IMA and 95-97.5% by ELISA. The use of serially samples increased the sensitivity of both tests and emphasizes the importance of re-testing individuals with a high suspicion of COVID-19.

The IMA had an overall sensitivity of 80% - considering all samples at different days from symptoms onset - which is more or less comparable to that of other point-of-care tests (POCs) commercially available, depending on days from debut of symptoms to test [13,27-30] but 100% for serial samples.

The advantage of POCs as screening tools is that they can reduce the amount of confirmatory testing required. We found that the positive predictive value (PPV) of all the methods used here (ELISA and IMA) is likely to be 93.5% or higher, given the high specificities observed (ranging from 95.0 to 100%). Therefore, individuals being tested positive, in a low-prevalence setting, are unlikely to require further confirmatory testing.

The sensitivity parameter of the IgM, IgA and IgG tests may vary according to the study design, a low negative predictive value (NPV) of a given test indicates that individuals testing negative but presenting clinical symptoms of COVID-19 need to be re-tested using another serological test or RT-PCR [27,31,32], Here, we found that the NPV of the IMA is likely to be comparable to that of a commercially available ELISA test (87-88%). The limitation of a given sample producing a false negative result is correlated to different factors, such as, time of testing in relation to symptom onset, changes in antibody levels during illness and severity of the disease. Several studies covering the use of ELISA, Chemiluminescence immunoassays (CLIA) and qualitative assays show that full diagnostic sensitivity for neither IgM nor IgG is reached before approximately 14-22 days from onset of symptoms. [29,30,33-35],

It has been reported that IgM detection was more variable than IgG, and detection was highest when IgM and IgG results were combined for both ELISA and POCs [27], The addition of IgA may improve sensitivity as it has been found to have higher titers than IgM [36].

Using IMA cartridges we observed a better performance of the IgA+IgM/IgG combination in terms of sensitivity while keeping the specificity at 100%.

Two previous studies have shown that antibodies to the nucleocapside antigen can be measured earlier than antibodies to the spike protein antigen [37,38]. In this study we included test against both antibodies but found little difference. An explanation for this may be that the samples were taken at different timepoints and only five study subjects had serial sampling performed.

Four of seven negative samples were taken less than 10 days before symptom onset. Studies have demonstrated that the median time for measurable antibodies following SARS-CoV-2 infection is five-seven days[ll,34], which possibly explains our result and emphasizes the importance of timely testing and re-testing.

The detailed clinical data including symptom onset and disease severity improved the interpretation of the results since antibody titers were found to be affected by both as previously reported [32,39]. Comparison to a well-tested commercial ELISA strengthens the evaluation of the novel IMA.

The current study lack sera of individuals infected with other coronaviruses to test for cross reactivity. A prior infection with other human coronaviruses may cause false positive results due to cross reactivity [32,40]. However, studies have indicated that cross reactivity mainly was detected for SARS-CoV-1 [40]. SARS-CoV-1 has never been diagnosed in Denmark. Negative control samples came from HIV positive individuals, which may have secondary hypogammaglobulinemia that potentially could lead to a falsely higher specificity. The analyses were performed on plasma and analysis on whole blood may decrease sensitivity of the point-of-care test (POC). Patients tested were all hospitalized, symptomatic and presented with moderate to severe disease. Studies have shown that the titers are higher in those with more severe symptoms [41]. Finally, the small number of individuals made it difficult to estimate the association between antibody response and disease severity.

In conclusion, our results show that the IMA IgM+IgA/IgG system is an effective supplemental diagnostic tool for COVID-19 with high sensitivity and specificity in hospitalized patients with moderate to severe disease. The test is rapid and can be performed at point-of-care as a supplement to RT-PCR in testing for active COVID-19 as well as a potential screening and testing tool for epidemiological studies in community settings enabling a rapid result without need for phlebotomy and handling of test tubes. Several lateral flow assays are already in use for COVID-19 IgM and IgG detection, but to our knowledge, this is the first semi-quantitative point of care assay with automized readout which measures IgA, IgM and IgG.

## Data Availability

The data are available and embedded in the paper. Raw data available upon request.

## Conflict of interest^1^, Funding^2^ and Corresponding author information^3^

^1^ P. Pah, A. De La C. Veras, M. Bade, J. Fock, S. and M. Donolato are employed at Blusense Diagnostics APS and have been part of the developing of ViroTrack Sero COVID IgM+lgA /

^2^This work was supported by BluSense Diagnostics in forms of providing the test kits for the study.

^3^ Maria Engel Moeller, Department of Infectious Diseases, Copenhagen University Hospital, Amager and Hvidovre Hospital, Hvidovre, Kettegaard Allé 30, 2650 Hvidovre, Denmark Telephone: +45 51345595

## Author contributions

Study planning: Marco Donolato, Jesper Eugen-Olsen, Thomas Benfield and Jeppe Fock. Sample collection: Maria Engel Moeller, Jesper Eugen-Olsen and Thomas Benfield.

Laboratory analysis: Pearlyn Pah, Melanie Bade, Antia De La Campa Veras. Manuscript drafting: Maria Engel Moeller, Simone Bastrup Israelsen, Thomas Benfield, Frederik N. Engsig and Jeppe Fock. Critical revision of manuscript: All authors approved the final manuscript.

## ACKNOWLEDGEMENTS

Authors acknowledge the work and contribution of all the health providers from Hvidovre Hospital.

## Supplementary Information

**Table S1:** Titers for both microfluidic quantitative immunomagnetic assay (IMA) (Virotrack) IgM+A and IgG and enzyme-linked immunosorbent assay (ELISA) IgA and IgG. Five patients (POS 1-5) had multiple blood-samples taken. For each patient, the use of supplementary oxygen and level of treatment is displayed.

|  | Age (10 year range) | Gender | Oxygen 1-14 L * | Oxygen 15-29 L * | Oxygen +30 L * | Limited treatment (%ICU)** | Limited treatment (%HLR)** | ICU admission*** | 30-days mortality | Days since symptoms | Virotrack |  |  |  | ELISA |  |  |  |
| --- | --- | --- | --- | --- | --- | --- | --- | --- | --- | --- | --- | --- | --- | --- | --- | --- | --- | --- |
|  |  |  |  |  |  |  |  |  |  |  | IgM+A |  | IgG |  | IgA |  | IgG |  |
| POS 01 | 70's | M |  |  | x |  |  |  |  | 23 | 1.4 | NEG | 2.3 | NEG | 0.5 | NEG | 0.4 | NEG |
|  |  |  |  |  |  |  |  |  |  | 29 | 11.8 | POS | 3.3 | NEG | 4.8 | POS | 3.8 | POS |
|  |  |  |  |  |  |  |  |  |  | 33 | 37.4 | POS | 24.7 | POS | 7.7 | POS | 6.0 | POS |
|  |  |  |  |  |  |  |  |  |  | 33 | 36.8 | POS | 15.6 | POS | 6.8 | POS | 5.6 | POS |
|  |  |  |  |  |  |  |  |  |  | 37 | 30.1 | POS | 30.8 | POS | 6.0 | POS | 6.4 | POS |
|  |  |  |  |  |  |  |  |  |  | 39 | 27.5 | POS | 7.0 | POS | 5.7 | POS | 2.1 | POS |
| POS 02 | 50's | M |  |  | x |  |  |  |  | 10 | 3.0 | NEG | 2.7 | NEG | 0.6 | NEG | 0.3 | NEG |
|  |  |  |  |  |  |  |  |  |  | 15 | 29.5 | POS | 47.4 | POS | 8.1 | POS | 5.1 | POS |
|  |  |  |  |  |  |  |  |  |  | 18 | 32.4 | POS | 25.7 | POS | 10.6 | POS | 7.4 | POS |
|  |  |  |  |  |  |  |  |  |  | 19 | 36.4 | POS | 36.3 | POS | 12.1 | POS | 7.6 | POS |
| POS 03 | 40's | M |  | x |  |  |  | x |  | 13 | 25.9 | POS | 41.8 | POS | 11.8 | POS | 4.2 | POS |
|  |  |  |  |  |  |  |  |  |  | 15 | 31.8 | POS | 27.3 | POS | 15.3 | POS | 6.5 | POS |
| POS 04 | 60's | F |  | x |  | x | x |  |  | 9 | 8.2 | POS | 26.8 | POS | 12.1 | POS | 1.9 | POS |
|  |  |  |  |  |  |  |  |  |  | 10 | 14.9 | POS | 28.0 | POS | 11.8 | POS | 3.4 | POS |
|  |  |  |  |  |  |  |  |  |  | 11 | 10.3 | POS | 36.6 | POS | 11.7 | POS | 4.3 | POS |
| POS 05 | 50's | M |  | x |  |  |  |  |  | 14 | 23.8 | POS | 18.0 | POS | 3.6 | POS | 1.5 | POS |
|  |  |  |  |  |  |  |  |  |  | 15 | 48.0 | POS | 23.8 | POS | 6.6 | POS | 3.5 | POS |
|  |  |  |  |  |  |  |  |  |  | 16 | 41.7 | POS | 45.1 | POS | 9.5 | POS | 5.3 | POS |
| POS 06 | 50's | F |  |  | x |  |  | x |  | 22 | 8.4 | POS | 55.7 | POS | 15.3 | POS | 8.5 | POS |
| POS 07 | 80's | F |  |  | x |  |  | x |  | 10 | 52.0 | POS | 51.4 | POS | 8.8 | POS | 5.1 | POS |
| POS 08 | 80's | F |  |  | x |  | x | x | x | 18 | 42.6 | POS | 6.3 | POS | 2.5 | POS | 2.7 | POS |
| POS 09 | 70's | F |  |  | x |  | x | x | x | 17 | 2.7 | NEG | 2.0 | NEG | 0.2 | NEG | 0.3 | NEG |
| POS 10 | 70's | M |  |  | x |  |  | x |  | 13 | 25.4 | POS | 80.6 | POS | 13.9 | POS | 8.8 | POS |
| POS 11 | 60's | F |  |  | x |  |  | x |  | 22 | 7.7 | POS | 6.4 | POS | 12.3 | POS | 7.0 | POS |
| POS 12 | 70's | M |  |  | x |  |  |  |  | 16 | 46.3 | POS | 11.8 | POS | 8.3 | POS | 7.3 | POS |
| POS 13 | 70's | M |  |  | x |  | x | x | x | 11 | 55.4 | POS | 38.5 | POS | 13.3 | POS | 6.1 | POS |
| POS 14 | 70's | M | x |  |  | x | x |  | x | 27 | 2.6 | NEG | 9.9 | POS | 6.1 | POS | 6.5 | POS |
| POS 15 | 80's | M | x |  |  | x | x |  |  | 14 | 32.4 | POS | 57.9 | POS | 11.5 | POS | 5.1 | POS |
| POS 16 | 70's | M |  |  | x |  |  | x |  | 47 | 5.0 | POS | 5.5 | POS | 11.6 | POS | 8.9 | POS |
| POS 17 | 60's | M |  |  | x |  |  |  |  | 29 | 14.7 | POS | 60.8 | POS | 11.0 | POS | 9.6 | POS |
| POS 18 | 20's | F |  | x |  |  |  |  |  | 8 | 1.4 | NEG | 1.6 | NEG | 0.6 | NEG | 0.4 | NEG |
| POS 19 | 70's | M |  | x |  | x | x |  |  | 27 | 10.0 | POS | 54.9 | POS | 10.9 | POS | 7.8 | POS |
| POS 20 | 50's | M |  |  | x |  |  |  |  | 12 | 17.9 | POS | 8.9 | POS | 2.5 | POS | 1.8 | POS |
| POS 21 | 70's | F |  |  |  | x | x |  |  | 1 | 3.1 | NEG | 2.6 | NEG | 1.0 | EQV | 0.4 | NEG |
| POS 22 | 80's | M | x |  |  | x | x |  |  |  | 1.6 | NEG | 1.4 | NEG | 0.2 | NEG | 0.3 | NEG |

\*Oxygen supply of either 1-14, 15-29 or more than 30 liters per minute
\*\* Limited treatment of either no resuscitation of the patient should cardiac arrest appear
(%HLR) or no admission to the intensive care unit (%ICU) or both.
\*\*\* Admission to the intensive care unit.

**Figure S1:**
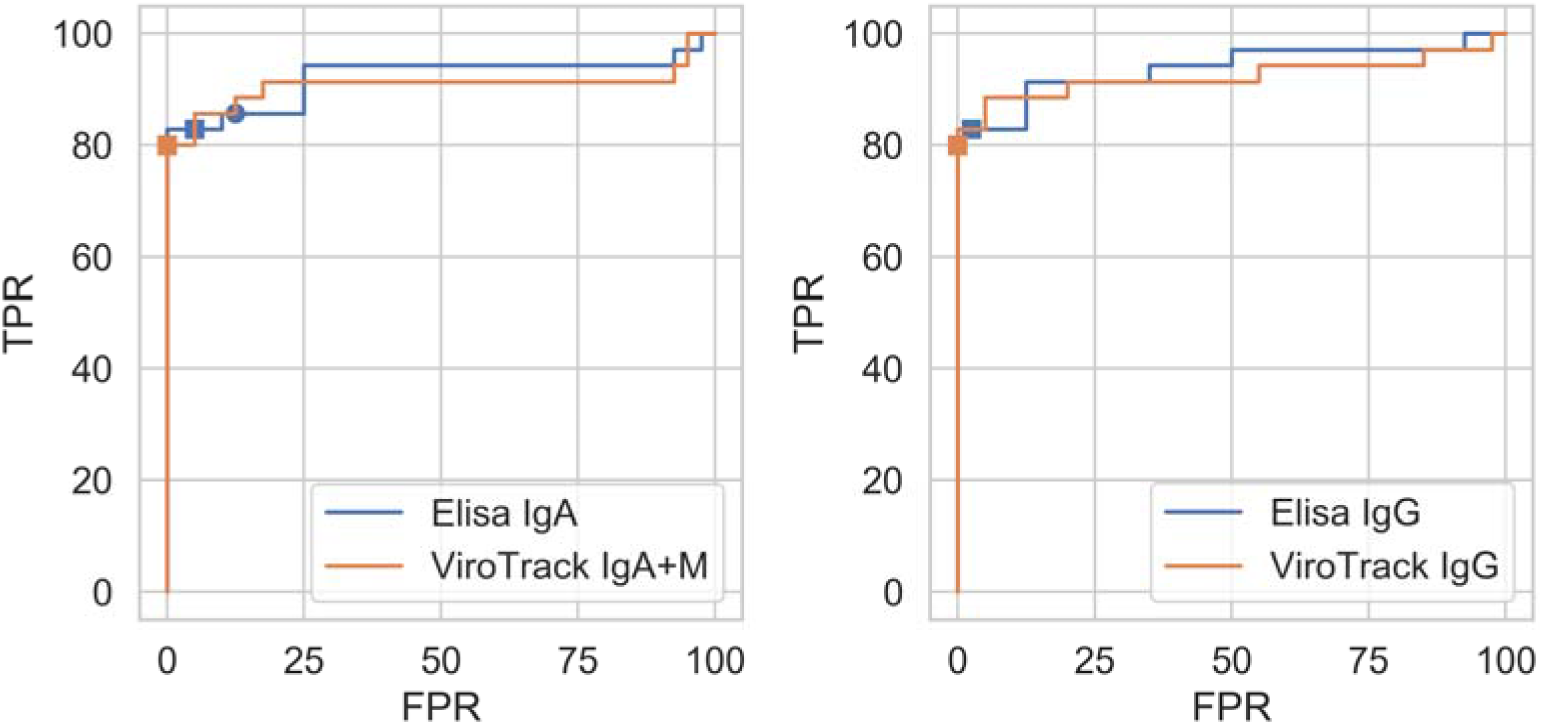
Receiver operating characteristic (ROC) curve of enzyme-linked immunosorbent assay [ELISA) and microfluidic quantitative immunomagnetic assays (IMA) (Virotrack). The area under curve (AUC) is as following: ELISA IgA (AUC 0.92), ELISA IgG (AUC 0.94), IMA IgA+IgM (AUC=0.91), IMA IgG (AUC=0.92). Square points correspond to the upper limit of the equivocal regions, and the circular points correspond to the lower limit of the equivocal regions.

**Table S2:** Confidence interval of analytical sensitivities, specificities, and predictive values. Equivocal/borderline results for enzyme-linked immunosorbent assay (ELISA) were treated as negatives (eqv. Neg) and as positives (eqv. Pos). No equivocal/borderline results were found in the microfluidic quantitative immunomagnetic assays (IMA) (Virotrack).

|  |  | **Sensitivity** | **Specificity** | **Agreement** | **PPV** | **NPV** |
| --- | --- | --- | --- | --- | --- | --- |
| **ELISA**<br>**(eqv. Neg)** | **IgA** | [66.4%;93.4%] | [83.1%;99.4%] | [80.1%;95.3%] | [78.6%;99.2%] | [72.6%;94.8%] |
|  | **IgG** | [66.4%;93.4%] | [86.8%;99.9%] | [81.7%;96.2%] | [82.8%;99.9%] | [73.2%;94.9%] |
|  | **IgA+IgG** | [66.4%;93.4%] | [83.1%;99.4%] | [80.1%;95.3%] | [78.6%;99.2%] | [72.6%;94.8%] |
| **ELISA**<br>**(eqv. Pos)** | **IgA** | [69.7%;95.2%] | [73.2%;95.8%] | [76.8%;93.4%] | [69.7%;95.2%] | [73.2%;95.8%] |
|  | **IgG** | [66.4%;93.4%] | [86.8%;99.9%] | [81.7%;96.2%] | [82.8%;99.9%] | [73.2%;94.9%] |
|  | **IgA+IgG** | [69.7%;95.2%] | [73.2%;95.8%] | [76.8%;93.4%] | [69.7%;95.2%] | [73.2%;95.8%] |
| **Virotrack** | **IgA+IgM** | [63.1%;91.6%] | [91.2%;100%] | [81.7%;96.2%] | [87.7%;100%] | [71.7%;93.8%] |
|  | **IgG** | [63.1%;91.6%] | [91.2%;100%] | [81.7%;96.2%] | [87.7%;100%] | [71.7%;93.8%] |
|  | **IgA+IgM+IgG** | [66.4%;93.4%] | [91.2%;100%] | [83.4%;97%] | [88.1%;100%] | [73.7%;95.1%] |

